# Meta-analysis reveals transcription factors and DNA binding domain variants associated with congenital heart defect and orofacial cleft

**DOI:** 10.1101/2025.01.30.25321274

**Authors:** Raehoon Jeong, Martha L. Bulyk

## Abstract

Many structural birth defect patients lack genetic diagnoses because there are many disease genes as yet to be discovered. We applied a gene burden test incorporating *de novo* predicted-loss-of-function (pLoF) and likely damaging missense variants together with inherited pLoF variants to a collection of congenital heart defect (CHD) and orofacial cleft (OC) parent-offspring trio cohorts (n = 3,835 and 1,844, respectively). We identified 17 novel candidate CHD genes and 10 novel candidate OC genes, of which many were known developmental disorder genes. Shorter genes were more powered in a “*de novo* only” analysis as compared to analysis including inherited pLoF variants. TFs were enriched among the significant genes; 14 and 8 transcription factor (TF) genes showed significant variant burden for CHD and OC, respectively. In total, 30 affected children had a *de novo* missense variant in a DNA binding domain of a known CHD, OC, and other developmental disorder TF genes. Our results suggest candidate pathogenic variants in CHD and OC and their potentially pleiotropic effects in other developmental disorders.

## Introduction

Various structural birth defects, ranging from congenital heart defect (CHD) to orofacial cleft (OC), affect approximately 3% of births each year in the United States (1) and account for about 20% of infant mortality (2). CHD patients have abnormalities in the structure of the heart at birth (3), while OC patients have an opening in their lips or palates (4). Improved understanding of their genetic etiology will improve the accuracy of genetic diagnoses and guide potential disease-specific treatment strategies.

Transcription factors (TFs) play key roles in orchestrating differentiation and establishing cell identity during development (5,6). Genetic variants that damage TF function can cause various developmental disorders (7). Sequence-specific TFs control gene expression programs by binding to recognition sites in the genome and regulating the expression of their target genes. Missense variants in the DNA binding domains (DBDs) of TFs can alter DNA binding activity and cause a wide range of diseases, including Mendelian diseases (8). For example, many of the pathogenic variants in *NKX2-5* and *TBX5* for CHD, and *IRF6* for OC, are found in their DNA binding domains (9,10). We thus hypothesized that DBD variants in other TF genes might also cause CHD or OC. Furthermore, we hypothesized that DBD variants not yet found to be pathogenic but that occur in TFs with DBD variants previously found to cause CHD or OC, might also cause CHD or OC.

Searching for genetic causes underlying structural birth defects requires genetic data from patients. In recent years, the Gabriella Miller Kids First pediatric research program (“Kids First” from here on) funded efforts to sequence the genomes of patients as well as the family trios. Such family trio studies have been a primary strategy to discover disease genes for structural birth defects (11–13). The trio design is crucial in detecting *de novo* variants in probands and ascertaining rare pathogenic variants, as demonstrated by the Deciphering Developmental Disorders (DDD) study (14). Most probands for CHD and OC are sporadic cases with unaffected parents (100% for CHD cohorts and 95.3% for OC cohorts in this study). Therefore, in this study, we searched for *de novo* variants and rare inherited variants in the probands.

The aim of our study was two-fold. First, we sought to discover novel disease genes in CHD and OC since more causal genes likely remain to be found (8,12,13,15). We boosted power to discover novel disease genes by combining data from multiple cohorts across the spectrum of syndromic and non-syndromic cases for CHD and OC, respectively (12,13,16–18). We utilized the PrimateAI variant effect prediction tool (19) to identify missense variants likely to be pathogenic more precisely than earlier studies (12,13). Furthermore, we applied the Transmission And *De novo* Association (TADA) (20) test to identify genes that show enrichment of putative damaging *de novo* inherited variants across different types of variant classes, such as missense and predicted loss-of-function (pLoF) variants (*i.e.*, nonsense, canonical splicing, and frameshift variants). This method has been successfully applied to discover potential autism genes (21).

Second, focusing on TFs because of their key roles in development and Mendelian diseases, we surveyed TFs and TF DBD variants for their potential association with CHD and OC. The resulting list of TFs and DBD variants are provided as a resource for future studies to evaluate whether they alter DNA binding activity (8,15).

## Results

### Genetic variants identified from multiple family trio cohorts of CHD and OC

To maximize power to discover novel disease genes, we combined genetic data from multiple CHD and, separately, OC cohorts. For CHD, we collected a non-redundant list of *de novo* variants and heterozygous predicted loss-of-function (pLoF) variants (*i.e.*, nonsense, canonical splicing, and frameshift variants) in probands from three prior studies (12,16,17), one of which is part of the Kids First program (17). In total, our list included variants from 3,835 family trios with a proband with CHD (**Supplementary Table 1**). For OC, we assembled genetic data from four Kids First cohorts (13,22) and the Deciphering Developmental Disorders (DDD) study (18), totaling 1,844 family trios (**Supplementary Table 1**). We combined those data with a list of *de novo* variants found in 757 family trios from Bishop *et al*. (11) and 603 family trios from Wilson *et al*. (18). For the Kids First cohort samples not analyzed in these two studies, we identified *de novo* variants from the whole-genome sequencing data using the slivar tool (23) (**Methods**).

### Missense variant effect prediction methods prioritized putatively damaging variants

Missense variant effect prediction methods aim to score missense variants according to their likelihood of being benign or pathogenic (24–32). Disease genes are expected to be enriched for damaging, and not neutral, variants. Therefore, we compared ten variant effect prediction tools in order to select one that best differentiates potentially damaging variants from neutral ones in the context of structural birth defects. For this, we scored *de novo* variants in known CHD genes (**Supplementary Table 2**) from CHD patients (12) (3,835 families with 113 variants) and unaffected siblings from an autism study (33) (2,179 families with 26 variants). We included unaffected siblings from an autism study because CHD cohorts did not have any genetic data from unaffected siblings and we can expect that unaffected siblings from an autism study likely did not have CHD diagnoses. Although these variants’ pathogenicity has not all been resolved, we nonetheless expect many of the *de novo* variants from CHD patients to be pathogenic and most of those from the unaffected children in the autism study to be benign for CHD.

We compared the performance of the ten tools in discriminating the two sets of variants at various score thresholds (**Figure 1A**). We aimed to select a method that highly enriches potentially pathogenic variants at the top quantile. Overall, PrimateAI (19) showed the highest area under the curve metric for both receiver operator characteristic (ROC) and precision-recall (**Supplementary Figure 1**). Although Missense Variant Pathogenicity (MVP) (25) performed similarly well, the number of variants from unaffected children that were falsely classified as pathogenic was higher than that using PrimateAI. For instance, there were 13 and 4 predicted pathogenic variants out of 26 *de novo* variants from unaffected children over the score percentile threshold of 0.75, using MVP and PrimateAI, respectively. Moreover, since PrimateAI does not use any disease association information in model training, we anticipate it is less likely to show overfitting. Therefore, we used PrimateAI to infer the likelihood of missense variant pathogenicity in all subsequent analyses in this study.

**Figure 1.**
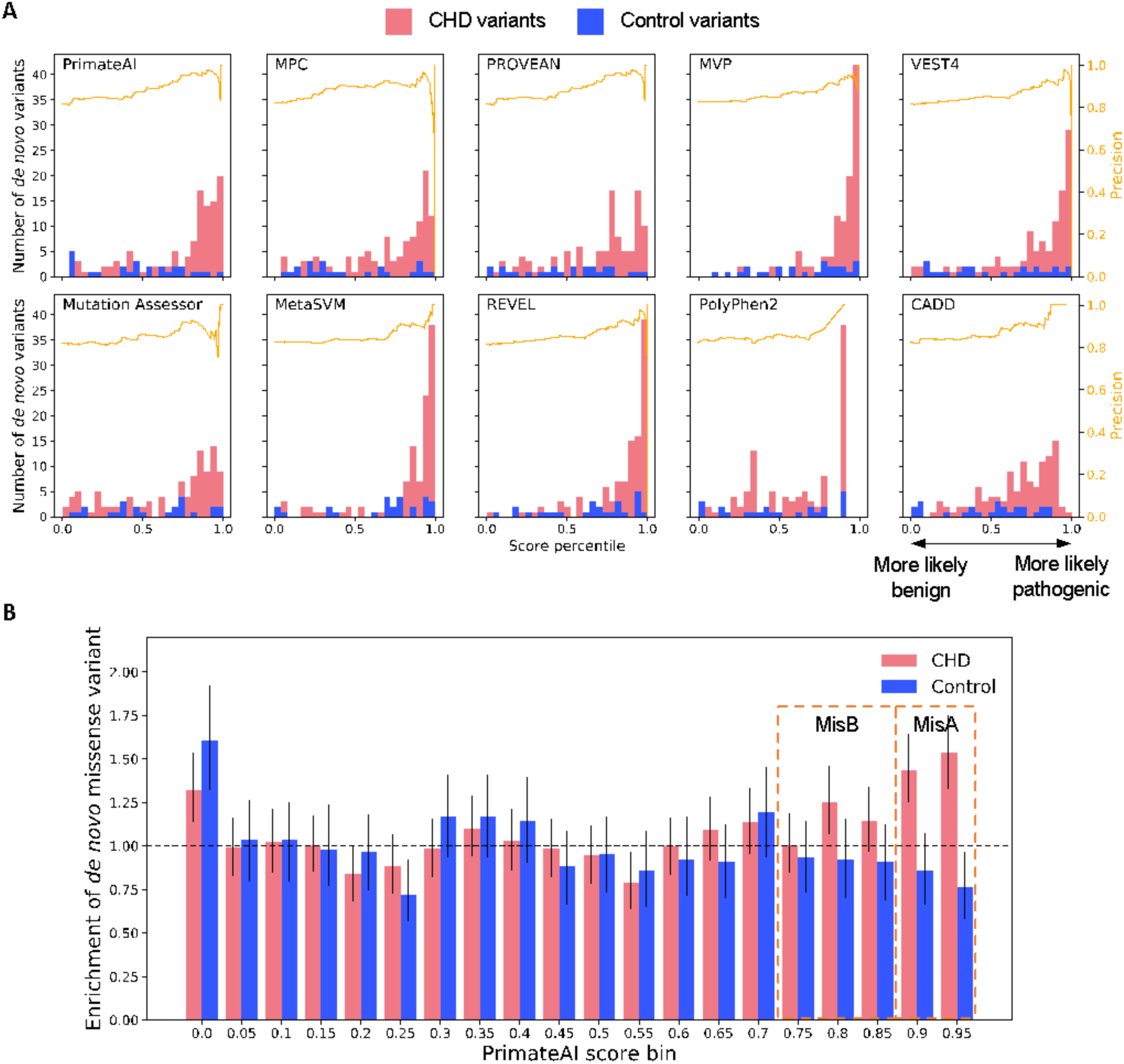
Comparison of missense variant prediction methods. (**A**) Number of variants in each score percentile bin, which corresponds to 5% increments, for ten missense variant effect predictions. Only *de novo* variants in 225 human CHD genes, which are listed in (**Supplementary Table 2**), are considered. The orange line depicts the precision at each percentile threshold. (**B**) Enrichment of missense variants in 5% PrimateAI score bins for all *de novo* variants in CHD patients and unaffected children. The error bars are 95% bootstrap confidence intervals. MisA, missense class A (PrimateAI ≥ 0.9); MisB, missense class B (0.75 < PrimateAI ≤ 0.9).

Next, we determined score thresholds to classify all *de novo* missense variants. Based on the total missense mutation rate (∼0.68 per generation), we inferred the expected number of *de novo* missense mutations in each 5% PrimateAI score bin. Then, we derived the enrichment of *de novo* missense variants in CHD versus control samples for each score bin **(Figure 1B**). The enrichment was more pronounced at the higher score bins. Therefore, we set two score thresholds: a stringent threshold of 0.9, and a more permissive, albeit still highly enriching, threshold of 0.75, to derive two groups of putatively damaging missense variants (PrimateAI ≥ 0.9 as MissenseA (MisA) and 0.75 ≤ PrimateAI < 0.9 as MissenseB (MisB)). These two subsets were enriched among CHD samples but depleted among control samples (**Supplementary Figure 2**). Variants with lower PrimateAI scores showed neither enrichment nor depletion in these samples. This is consistent with enrichment of *de novo* missense variants predicted to be damaging in patients of CHD and autism (11,33). From here on, we considered *de novo* and inherited pLoF, *de novo* MisA, and *de novo* MisB variants as putatively damaging. We used the same score thresholds for the analysis of the OC patient cohorts.

### Detection of genes with enrichment of putatively damaging *de novo* and rare variants

Next, to identify candidate CHD and OC genes, we analyzed the *de novo* pLoF, MisA, and MisB variants and rare inherited pLoF variants using the transmission and *de novo* association (TADA) model (20). This model integrates enrichment of *de novo* variants based on a mutational model (34) and the enrichment of variants from cases compared to those from controls. The test calculates a Bayes factor that captures the enrichment of putatively damaging variants of different types. We considered 3,578 unaffected parents in an autism cohort as controls (12,35).

We detected 46 and 22 significant genes (q value < 0.1) for CHD and OC, respectively, of which some are known CHD or OC genes (**Supplementary Tables 2-5**). Since genes with no depletion of pLoF variants in a healthy population are not likely to be structural birth defect genes, we excluded genes with a gnomAD (36) loss-of-function observed/expected upper bound fraction (LOEUF) > 1. Most candidate genes had both pLoF and missense variants contributing to the enrichment (**Figure 2**). Thus, integrating the variant types was useful in detecting candidate disease genes.

**Figure 2.**
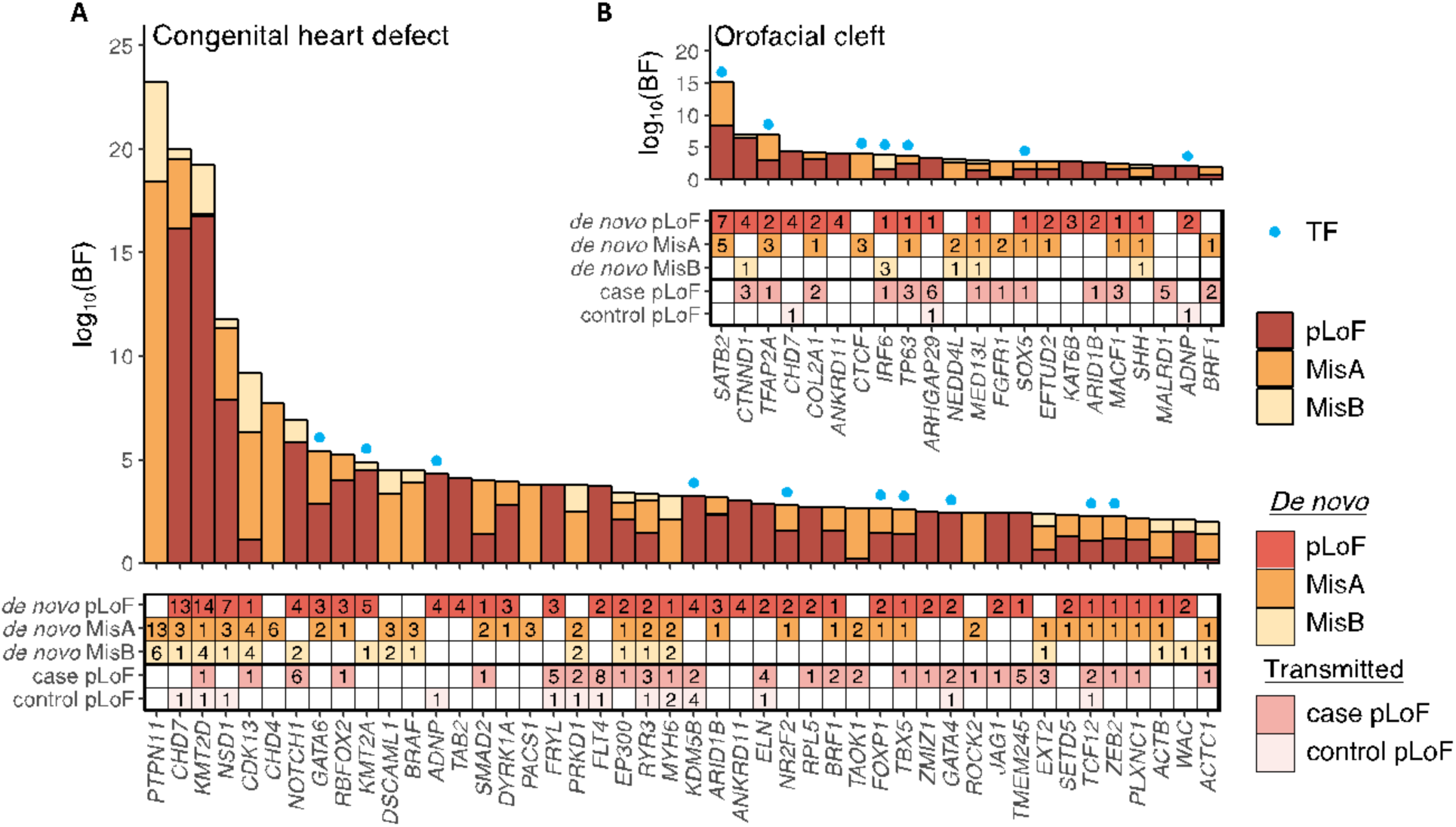
Bayes factor for each variant type’s enrichment in candidate disease genes. (A and. **B**) (Top) Bayes factor contribution by MisA, MisB, and pLoF variants in TADA for **(A)** CHD and **(B)** OC in the “*de novo* + case/control” setting. Only positive Bayes factor contributions in candidate genes (*q* value < 0.1) with LOEUF < 1 are displayed (CHD: 46 genes, OC: 22 genes). (Bottom) Number of variants in each category. BF, Bayes factor; TF, transcription factor.

17 of the 46 genes identified in the CHD analysis cohorts were not known CHD genes (*i.e.*, not significant in studies of individual cohorts and not annotated as CHD genes). 8 of the 22 genes identified in the OC analysis cohorts were not known OC genes; known OC genes were taken from the Genomics England PanelApp (37) ‘Clefting’ version 4.0 list. CHD and OC patients are at higher risk for other congenital anomalies (38,39). Indeed, several of these genes are developmental disorder genes, such as *TAOK1*, *WAC*, *PACS1*, *FOXP1*, *BRAF*, *SETD5*, and *ZMIZ1* (phenotype MIM numbers: 619575, 616708, 615009, 613670, 613706, 615761, and 618659, respectively). In a recent study on CHD (40), a *de novo* variant in *SETD5* was considered to be a positive diagnosis. However, that study did not perform an enrichment analysis to identify novel disease genes. Similarly, 7 of the 8 novel candidate OC genes – *MED13L*, *SOX5*, *KAT6B*, *ARID1B*, *MACF1*, *ADNP*, and *BRF1* – are linked to various developmental disorders (phenotype MIM numbers: 6616789, 616803, 616170, 135900, 618325, 615873, and 616202, respectively). These results are consistent with the known associations of CHD and OC with neurodevelopmental disorders (41,42).

More than half of the significant genes in CHD and OC showed probands with an inherited pLoF variant in the candidate disease gene (27 out of 46 for CHD and 13 out of 22 for OC). Two of the OC family trios (one with a *CTNND1* pLoF variant and another with an *AFHGAP29* pLoF variant) had an affected parent who passed on the pLoF variant. However, most inherited pLoF variants in candidate and known disease genes were inherited from unaffected parents, suggesting the possibility of incomplete penetrance.

### *De novo* missense variants in CHD and OC genes

Predicting the pathogenic effects of missense variants is challenging, and many are classified as variants of uncertain significance (VUSs) in ClinVar (43). Although we selected PrimateAI for this study, predictions by other methods can also be informative. As a resource for clinical researchers, we provide a table of predictions for the *de novo* missense variants identified in CHD and OC genes (**Supplementary Tables 6 and 7**). These tables include *de novo* missense variants in known CHD or OC genes (**Supplementary Table 2** and **3**) and candidate CHD or OC genes in the respective cohorts. In addition to scores from the tools we compared in **Figure 1**, we also include scores from the more recent AlphaMissense tool (44).

### Coding sequence length affects which TADA model detects enrichment in a gene

To evaluate the utility of incorporating inherited pLoF variants in the case/control setting (*i.e.,* “*de novo* & case/control”), we compared against the enrichment obtained using just *de novo* variants with TADA (*i.e.*, “*de novo* only”). Surprisingly, using just the *de novo* variants yielded more candidate CHD genes (**Supplementary Table 4**) than using the “*de novo* & case/control” setting; 24 and 10 genes were exclusively significant in “*de novo* only” and “*de novo* & case/control” settings, respectively. The 24 genes that were significant (*i.e.*, TADA *q* value < 0.1 and LOEUF < 1) only in the “*de novo* only” setting had no rare inherited pLoF variants in the cohorts, which lowered the Bayes factor estimates when case/control data were incorporated. Since approximately 90% of these genes are highly constrained with LOEUF < 0.3 (*i.e.*, in approximately the top 10% of all protein-coding genes), pLoF variants in these genes are expected to be extremely rare in unaffected individuals. Since longer genes are expected to have more pLoF variants on average, we compared the lengths of genes unique to each setting. The coding sequence lengths of the 10 genes that were uniquely significant in the “*de novo* & case/control” model were significantly longer than those of the 24 genes uniquely significant in the “*de novo* only” model (*p* = 0.019, one-sided Wilcoxon rank-sum test; **Figure 3**). The LOEUF estimates of genes in the two sets were not significantly different (*P* > 0.05, Wilcoxon rank-sum test). We observed similar trends for candidate OC genes (**Supplementary Table 5** and **Supplementary Figure 3**). Altogether, these results demonstrate that the coding sequence length of genes affects their identification as significant disease genes by the “*de novo* only” versus the “*de novo* & case/control” TADA model. This effect is likely because longer genes have a greater chance that pLoF variants are present in a population and inherited, thereby contributing to increased enrichment in the “*de novo* & case/control” setting; in contrast, shorter genes have a lower expected mutation rate for pLoF variants, thus each *de novo* variant contributes to a greater amount of enrichment.

**Figure 3.**
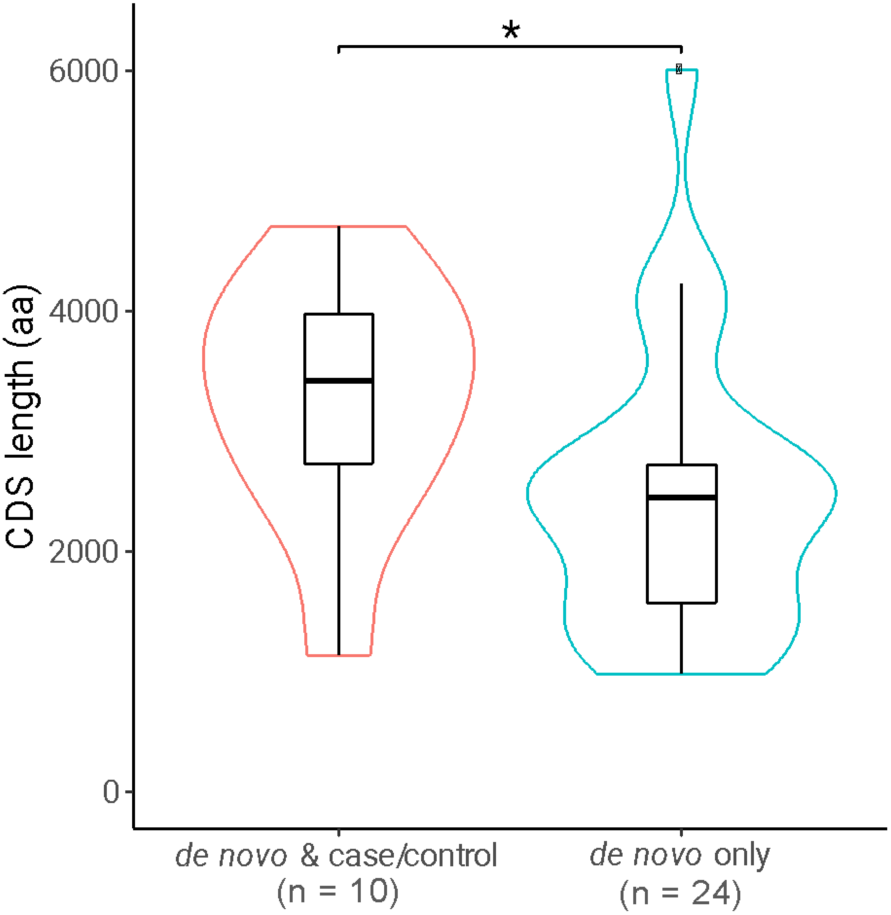
Coding sequence length of significant CHD genes by discovery model. Distribution of coding sequence length for the significant genes unique to the “*de novo* & case/control” model and “*de novo* only” model. The number of genes is labeled below each category. CDS, coding sequence; aa, amino acid. * *P* < 0.05, one-sided Wilcoxon rank-sum test.

### TF DBD variants identified in candidate CHD and OC disease genes

Because of the known role of TFs in CHD (45) and OC (46), we examined how many significant genes from our analysis were TFs (47). For CHD, there were 14 TFs that showed significant enrichment in either “*de novo* & case/control” or “*de novo* only” analysis (**Table 1**, **Figure 2**). For OC, 7 TFs showed significant enrichment (**Table 1**, **Figure 2**). For both CHD and OC, TFs were significantly enriched among the significant genes (*p* = 0.006 and *p* = 0.016, respectively, one-sided Fisher’s exact test).

**Table 1.** Transcription factors significantly enriched for predicted deleterious *de novo* variants. LOEUF, loss-of-function observed/expected upper bound fraction (36); pLoF, predicted loss-of-function; MisA, PrimateAI > 0.9; MisB, PrimateAI 0.75-0.9. ^a^ novel candidate CHD genes. ^b^ novel candidate OC genes.

| Gene | LOEUF | de novo variants |  |  | Inherited variants |  | de novo & case/control q value | de novo only q value |
| --- | --- | --- | --- | --- | --- | --- | --- | --- |
|  |  | pLoF | MisA | MisB | Case pLoF | Control pLoF |  |  |
| Congenital heart defect |  |  |  |  |  |  |  |  |
| GATA6 | 0.174 | 3 | 2 | 0 | 0 | 0 | 2.5×10<sup>-5</sup> | 3.5×10<sup>-6</sup> |
| KMT2A | 0.065 | 5 | 0 | 1 | 0 | 0 | 3.2×10<sup>-4</sup> | 5.0×10<sup>-5</sup> |
| ADNP | 0.123 | 4 | 0 | 0 | 0 | 1 | 5.8×10<sup>-4</sup> | 3.1×10<sup>-4</sup> |
| KDM5B<sup>a</sup> | 0.572 | 4 | 0 | 0 | 2 | 4 | 0.012159 | 5.2×10<sup>-4</sup> |
| NR2F2 | 0.217 | 2 | 1 | 0 | 0 | 0 | 0.014122 | 0.002103 |
| FOXP1<sup>a</sup> | 0.175 | 2 | 1 | 0 | 0 | 0 | 0.029404 | 0.003100 |
| TBX5 | 0.135 | 1 | 1 | 0 | 1 | 0 | 0.031389 | 0.053298 |
| GATA4 | 0.527 | 2 | 0 | 0 | 2 | 1 | 0.040077 | 0.050796 |
| TCF12 | 0.372 | 1 | 1 | 0 | 2 | 1 | 0.051542 | 0.068473 |
| ZEB2 | 0.107 | 1 | 1 | 0 | 1 | 0 | 0.058741 | 0.131609 |
| KLF2<sup>a</sup> | 0.710 | 1 | 1 | 0 | 0 | 0 | 0.204573 | 0.032261 |
| SMAD4 | 0.222 | 0 | 2 | 0 | 0 | 0 | 0.209108 | 0.035057 |
| MEIS2<sup>a</sup> | 0.184 | 2 | 0 | 0 | 0 | 0 | 0.271015 | 0.055872 |
| CTCF<sup>a</sup> | 0.148 | 0 | 2 | 0 | 0 | 0 | 0.278293 | 0.058374 |
| Orofacial cleft |  |  |  |  |  |  |  |  |
| SATB2 | 0.091 | 7 | 5 | 0 | 0 | 0 | 3.86×10<sup>-14</sup> | 5.77×10<sup>-15</sup> |
| TFAP2A | 0.261 | 2 | 3 | 0 | 1 | 0 | 2.84×10<sup>-6</sup> | 7.70×10<sup>-6</sup> |
| CTCF<sup>b</sup> | 0.148 | 0 | 3 | 0 | 0 | 0 | 0.011737 | 0.001484 |
| IRF6 | 0.132 | 1 | 0 | 3 | 1 | 0 | 0.002951 | 0.007637 |
| TP63 | 0.267 | 1 | 1 | 0 | 3 | 0 | 0.003631 | 0.072430 |
| SOX5<sup>b</sup> | 0.188 | 1 | 1 | 0 | 1 | 0 | 0.018728 | 0.058691 |
| ADNP<sup>b</sup> | 0.123 | 2 | 0 | 0 | 0 | 1 | 0.092444 | 0.088307 |
| GRHL2<sup>b</sup> | 0.270 | 2 | 0 | 0 | 0 | 0 | 0.328840 | 0.076571 |

There were 5 and 3 candidate CHD and OC TF genes, respectively, that are not yet established CHD or OC disease genes. For CHD, we identified *KDM5B*, *FOXP1*, *KLF2*, *MEIS2*, and *CTCF*. For OC, we identified *SOX5, ADNP,* and *GRHL2*. Two candidate CHD TF genes – *KDM5B* and *FOXP1* – were also statistically implicated in a similar CHD study (48) that aggregated *de novo* variants from two (12,16) of the 3 studies that we analyzed. Nevertheless, *KDM5B*, *FOXP1*, *MEIS2*, and *CTCF* are known developmental disorder genes (phenotype MIM numbers: 618109, 613670, 600987, and 615502, respectively). Some children with mutations in these genes have been reported to show heart defects (49–52). *KLF2* has not been directly associated with CHD, but its zebrafish homologue *klf2* is required for heart valve formation (53). A non-coding variant that causes over-expression of *Grhl2* in mice led to orofacial cleft phenotypes (54).

Since DNA binding activity plays a crucial role in TF function, we searched for TF DBD missense variants in known developmental disorder genes. We developed a pipeline to filter for missense variants in the TF DBDs based on a set of 62 DBD classes in the Pfam database (55) (**Supplementary Table 8**) and the protein domain prediction model HMMer (56). Without filtering for disease genes, there were 46 and 11 *de novo* TF DBD missense variants in the CHD and OC cohorts, respectively (**Supplementary Table 9**); with filtering, there were 17 and 13 DBD missense variants, respectively (**Table 2**). Some of these variants are in CHD, OC, and other developmental disorder genes that are mostly haploinsufficient, characterized by low LOEUF estimates (**Table 2**). Based on PrimateAI, they were all predicted to be pathogenic (PrimateAI rank score > 0.8). We hypothesize that these variants damage the TFs’ DNA binding activity.

**Table 2.** *De novo* TF DBD missense variants in genes associated with CHD, OC, or developmental disorder genes. The table lists *de novo* TF DBD variants from our analysis in genes that are either significantly enriched in our study (marked with an asterisk [*]) or are reported as CHD, OC, or developmental disorder genes. For developmental disorders, the specific syndrome is written in parentheses. PrimateAI rank score is a percentile score (range 0-1) based on the raw PrimateAI score. CAKUT, congenital anomalies of kidney and urinary tract; CDH, congenital diaphragmatic hernia; ETS, erythroblast transformation specific; IRF, interferon regulatory factor; AP-2, activator protein 2; EEC, Ectrodactyly, ectodermal dysplasia, and cleft lip/palate. ^a^ Candidate CHD gene based on damaging variant enrichment. * Significant enrichment of damaging variants in this study.

| Developmental disorder | Gene | LOEUF | Amino acid change | PrimateAI rank score | Variant | DBD (Pfam ID) |
| --- | --- | --- | --- | --- | --- | --- |
| **Congenital heart defect** |  |  |  |  |  |  |
| CHD | *FOXP1* * | 0.175 | F499L | 0.99469 | 3:70976974:A:T | Forkhead (PF00250) |
| CHD (Axenfeld-Rieger syndrome) | *FOXC1* | 0.311 | T88I | 0.94564 | 6:1610708:C:T | Forkhead (PF00250) |
| CHD (Wiedemann-Steiner syndrome) | *KMT2A* | 0.065 | K1186E | 0.87072 | 11:118478188:A:G | CXXC zinc finger (PF02008) |
| CHD (Holt-Oram syndrome) | *TBX5* * | 0.135 | I227T | 0.98142 | 12:114385551:A:G | T-box (PF00907) |
| CHD | *TCF12* * | 0.372 | H631Q | 0.98114 | 15:57273177:C:G | Helix-loop-helix (PF00010) |
| CHD | *NR2F2* * | 0.217 | C96F | 0.98292 | 15:96332392:G:T | C4 zinc finger (PF00105) |
| CHD | *GATA6* * | 0.174 | R456G | 0.90881 | 18:22181516:C:G | GATA zinc finger (PF00320) |
| CHD | *GATA6* * | 0.174 | R456H | 0.92717 | 18:22181517:G:A | GATA zinc finger (PF00320) |
| CHD<sup>a</sup> | *KLF2* * | 0.71 | C334Y | 0.99874 | 19:16326964:G:A | C2H2 zinc finger (PF00096) |
| CHD (DiGeorge syndrome) | *TBX1* | 0.427 | L293F | 0.98054 | 22:19765767:C:T | T-box (PF00907) |
| CAKUT | *PBX1* | 0.255 | R235Q | 0.95192 | 1:164807544:G:A | Homeodomain (PF00046) |
| CAKUT | *TBX18* | 0.193 | T305A | 0.81286 | 6:84747946:T:C | T-box (PF00907) |
| CDH (Cardiac-urogenital syndrome) | *MYRF* | 0.117 | Q403H | 0.8663 | 11:61774060:G:C | NDT80 / PhoG (PF05224) |
| CDH (Cardiac-urogenital syndrome) | *MYRF* | 0.117 | L479V | 0.86641 | 11:61776368:C:G | NDT80 / PhoG (PF05224) |
| Den Hoed-de Boer-Voisin syndrome | *SATB1* | 0.293 | E547K | 0.96969 | 3:18352132:C:T | CUT (PF02376) |
| Speech language disorder | *FOXP2* | 0.219 | R553H | 0.9789 | 7:114662075:G:A | Forkhead (PF00250) |
| Craniosynostosis | *ERF* * | 0.261 | K96N | 0.96845 | 19:42249912:C:A | ETS (PF00178) |
| **Orofacial cleft** |  |  |  |  |  |  |
| OC (van der Woude syndrome) | *IRF6\** | 0.132 | N88D | 0.86413 | 1:209796465:T:C | IRF (PF00605) |
| OC (van der Woude syndrome) | *IRF6\** | 0.132 | R84H | 0.84067 | 1:209796476:C:T | IRF (PF00605) |
| OC (Glass syndrome) | *SATB2\** | 0.091 | R667G | 0.94148 | 2:199272414:G:C | Homeodomain (PF00046) |
| OC (Glass syndrome) | *SATB2\** | 0.091 | R399H | 0.90829 | 2:199328888:C:T | CUT (PF02376) |
| OC (Glass syndrome) | *SATB2\** | 0.091 | L394S | 0.95427 | 2:199328903:A:G | CUT (PF02376) |
| OC (Glass syndrome) | *SATB2\** | 0.091 | R389L | 0.9951 | 2:199348708:C:A | CUT (PF02376) |
| OC (Glass syndrome) | *SATB2\** | 0.091 | R389C | 0.99811 | 2:199348709:G:A | CUT (PF02376) |
| Lamb-Shaffer syndrome | *SOX5* | 0.188 | H582Y | 0.9764 | 12:23543238:G:A | HMG_box (PF00505) |
| OC | *TFAP2A\** | 0.261 | R256Q | 0.921 | 6:10404511:C:T | AP-2 (PF03299) |
| OC | *TFAP2A\** | 0.261 | S249L | 0.98055 | 6:10404532:G:A | AP-2 (PF03299) |
| OC (EEC syndrome) | *TP63* | 0.267 | C347F | 0.94957 | 3:189868627:G:T | P53 (PF00870) |
| Holoprosencephaly | *SIX3* | 0.323 | W253R | 0.99697 | 2:44942861:T:A | Homeodomain (PF00046) |
| Ayme-Gripp syndrome | *MAF* | 0.537 | R294W | 0.99834 | 16:79599023:G:A | bZIP_MAF (PF03131) |

## Discussion

We aggregated multiple parent-offspring trio cohorts of CHD and OC to detect 46 and 22 genes, respectively, with enrichment of damaging *de novo* variants and inherited pLoF variants. Of those, 17 were novel candidate CHD genes and 10 were novel candidate OC genes (**Supplementary Tables 3 and 4**). Further studies are needed to validate which of these are true disease genes for CHD and OC. Increasing the sample sizes of family trio cohorts will be key to discovering more candidate disease genes; however, thousands of family trios are still insufficient to discover most of the disease genes. As there are likely hundreds of genes causing these structural birth defects, the likelihood of observing multiple cases with damaging *de novo* variants in the same gene is still low. Kaplanis and colleagues estimated that sequencing hundreds of thousands of parent-offspring trios will be necessary to reach sufficient power to detect about 80% of developmental disorder genes based on analysis of *de novo* variants (14).

We evaluated the performance of multiple missense variant effect prediction methods to prioritize candidate pathogenic variants. While most methods were able to discriminate *de novo* missense variants in CHD genes found in CHD patients from those found in unaffected children, PrimateAI was the most effective and led to the identification of more *de novo* missense variants. We also provide a list of *de novo* missense variants in known and candidate CHD and OC genes as a resource (**Supplementary Tables 6 and 7**).

Incorporating the number of inherited pLoF variants in cases and controls into enrichment analyses led to some significant genes not reaching significance with *de novo* variants alone. However, in the current sample size, there were many genes with no inherited pLoF variants, and many of them were only significant in the “*de novo* only” analysis. These genes were generally shorter than the genes identified uniquely by the “*de novo* & case/control” analysis, suggesting that gene length affects which model may be better powered. Moreover, applying both the “*de novo* only” and the “*de novo* & case/control” model is useful for detecting as many candidate disease genes as possible.

In this study, we analyzed only pLoF and missense variants. Copy number variations (CNVs) that increase or decrease gene dosage also play a role in structural birth defects (57). Therefore, calling *de novo* and inherited CNVs in the affected children and testing their enrichment in individual genes will increase the chance of disease gene discovery in future studies (21). In terms of inherited variants, we considered only pLoF variants because the effects of missense variants are more difficult to predict. Including inherited missense variants in the model may potentially increase power, but ensuring high precision in pathogenicity prediction will be essential. TFs were enriched among the identified genes. We identified many *de novo* TF DBD missense variants in genes that were significantly enriched in CHD or OC or that are known CHD, OC, or developmental disorder genes. The identified variants were predicted to be pathogenic by PrimateAI. Some of the TFs with TF DBD variants in the CHD cohort are known to cause other developmental disorders, such as congenital diaphragmatic hernia and congenital anomalies of kidneys and urinary tract (58,59). These results suggest that these TFs are pleiotropic and that other mutations in them may cause heart defects in some patients.

Variant effect prediction tools are only moderately accurate, at best, in distinguishing TF DBD missense variants with altered DNA binding activity (15). Future studies using DNA binding assays, such as protein binding microarrays (PBMs) (8,60), will be needed to determine which of the identified CHD and OC variants alter DNA binding activity and in what manner they do so.

## Methods

### Genetic data from family trio cohorts of CHD and OC

We aggregated multiple datasets to maximize statistical power to detect disease genes. For CHD, we downloaded *de novo* variant data from two exome-sequencing studies (12,16) and one genome-sequencing study (17). We also downloaded the list of rare inherited pLoF variants from Jin *et al*. (12). We identified overlapping samples by comparing the set of *de novo* variants from each proband. After removing duplicate samples, there were a total of 3,835 unique family trios.

For OC, we downloaded genotype data from 4 cohorts from the Gabriella Miller Kids First data portal (61). Their database of Genotypes and Phenotypes (dbGaP) IDs were phs001168 (n = 376 trios), phs001997 (n = 404 trios), phs001420 (n = 262 trios), and phs002595 (n = 351 trios). In addition, we downloaded a list of *de novo* variants from 374 European (phs001168), 267 Colombian (phs001420), and 116 Taiwanese (phs001997) family trios from Table S3 of Bishop *et al*. (13). We also downloaded a list of *de novo* variants from 603 family trios from Table S4 of Wilson *et al*. (18). We downloaded *de novo* variant data from unaffected siblings in families in an autism cohort (33) to compare variant enrichment statistics. Lastly, we downloaded heterozygous pLoF variants from 3,578 unaffected parents in an autism cohort as controls (12,35). We analyzed all genetic variants based on the GRCh38 human reference genome. The downloaded variants in hg19 were lifted over to the GRCh38 human reference. We performed variant calling and curation just for the 484 OC samples not included in Bishop *et al*. (13).

### Identifying *de novo* variants and rare inherited variants in the OC cohorts

For the samples not included in Bishop *et al*. (13) (n = 484), we applied different strategies for identifying *de novo* predicted-loss-of-function (pLoF) and missense variants. pLoF variants consist of nonsense, splice site, and frameshift variants. Since trio-based variant calls (*i.e.*, vcf files) provided in the Gabriella Miller Kids First data portal (61) showed false negatives in *de novo* single nucleotide variants (SNVs), we derived *de novo* SNVs based on the gvcf files of the three family members in each trio.

For SNVs, which span pLoF and missense variants, we identified *de novo* variants by 1) merging gvcf files of the three family members in each trio using GLNexus (62) with the ‘gatk’ setting and 2) using slivar (23) to filter for variants that are heterozygous in the proband but homozygous reference in the two parents. We further filtered for those with the maximum population allele frequency in gnomAD (36) of less than 5×10^-5^, no homozygous individuals in gnomAD, and TOPMed (63) allele frequency of less than 5×10^-5^.

In contrast, we used *de novo* insertions and deletions (indels) identified in the trio-based variant calls. For indel pLoF variants, we 1) downloaded the family-based vcf files from the Gabriella Miller Kids First data portal and 2) filtered for variants that are heterozygous in the proband but homozygous reference in the two parents using slivar (23). The variants were filtered for having genotype quality (GQ) greater than 20 and read depth (DP) greater than 6. We also filtered for those with a maximum population allele frequency in gnomAD (36) of less than 5×10^-5^, no homozygous individuals in gnomAD, and TOPMed (63) allele frequency of less than 5×10^-5^.

For all OC samples, we identified rare inherited pLoF variants by filtering for variants with a heterozygous genotype in the proband and only one parent with a heterozygous genotype using the family-based vcf files from the Gabriella Miller Kids First data portal. We also filtered for those with the maximum population allele frequency in gnomAD (36) of less than 5×10^-5^, no homozygous individuals in gnomAD, and TOPMed (63) allele frequency of less than 5×10^-5^.

### Comparison of missense variant effect prediction methods

We compared the performance of ten missense variant effect prediction methods: PrimateAI (19), MPC (31), PROVEAN (26), MVP (25), VEST4 (30), MutationAssessor (32), MetaSVM (28), REVEL (29), PolyPhen2 (24), and CADD (27). These tools’ scores for missense variants were accessed from the database for nonsynonymous SNPs’ functional predictions (dbNSFP) version 4.5 (64). To compare between scores easily, we utilized the rank scores, which range from 0 to 1 and correspond to the percentile among missense variants. We compared their performance in discriminating *de novo* missense variants in CHD genes (**Supplementary Table 2**) from CHD patients from those from unaffected children. There were a total of 3,836 CHD family trios (12,16,17) and 2,179 control family trios (33) that carried 113 and 26 *de novo* variants in CHD genes, respectively. We computed their area under the curve for receiver operator characteristic (ROC) and precision-recall to compare their performance.

Next, we determined the appropriate PrimateAI score thresholds for potentially damaging variants. Across all genes, we estimated the enrichment of *de novo* missense variants for CHD families and control families in each of the 5% score bins. The expected number of *de novo* missense variants per family was the sum of all missense mutation rates (∼ 0.68 per generation). Then, we bootstrapped sampled CHD and control families to establish the respective 95% confidence intervals of the enrichment estimates. Ultimately, based on **Figure 1B**, we selected PrimateAI ≥ 0.9 and 0.75 ≤ PrimateAI < 0.9 as the two missense variant groups – MisA and MisB.

### Testing enrichment of damaging *de novo* and rare inherited variants

We used the TADA model (20) to detect genes with an enrichment of potentially damaging variants (*i.e.* predicted-loss-of-function (pLoF), missense with PrimateAI (19) rank score ≥ 0.9 (MisA), or missense with PrimateAI rank score 0.75∼0.9 (MisB)) from the number of *de novo* variants and mutation rate estimates. We derived the per-gene mutation rates for MisA, MisB, and pLoF based on estimates in Samocha *et al.* (34) and gnomAD (36). We multiplied the per-gene missense mutation rate *μ*_Mis, gene_ by 0.1 and 0.15, to derive *μ*_MisA, gene_ and *μ*_MisB, gene_, respectively, as all possible MisA and MisB variants are expected be 0.1 and 0.15 of all missense variants. We added the per-gene nonsense, splice site and frameshift mutation rates to derive the per-gene pLoF mutation rates.

We applied TADA to 17,488 autosomal genes with LOEUF estimates in gnomAD (36). We performed the test once including inherited pLoF variants and once without to compare the effect of inherited variants. Multiple hypothesis correction across all genes was applied using the *q* value estimates. We considered genes with *q* value < 0.1 and gnomAD’s LOEUF < 1 to be significant. We excluded genes with LOEUF ≥ 1 because it suggests that there is negligible selective constraint against predicted-loss-of-function variants in those genes.

### Identifying TF DBD variants in candidate disease genes

We identified disease-associated TF genes based on a list of 1,639 TFs (47). Then, we determined the location of the DBDs using a set of 62 DBD classes in the Pfam database version 35.0 (55) (**Supplementary Table 5**) and the protein domain prediction model HMMer (56). We considered only canonical transcripts and amino acid sequences based on GENCODE (65) in annotating whether the missense variants fall within a DBD.

## Data availability

For CHD, we downloaded *de novo* variant data from two exome-sequencing studies (12,16) and one genome-sequencing study (17). We also downloaded the list of rare inherited pLoF variants from Jin *et al*. (12). For OC, we downloaded genotype data from 4 cohorts from the Gabriella Miller Kids First data portal (61). Their database of Genotypes and Phenotypes (dbGaP) IDs were phs001168, phs001997, phs001420, and phs002595.

## Code availability

Code and data for generating the figures is available at https://github.com/BulykLab/CHD-OC-manuscript-figures.

## Acknowledgments

We thank members of the Bulyk lab for helpful discussion. This work was funded by NIH grants R03 HD099358 and R01 HG010501 to M.L.B.

## Author Contributions

R.J. and M.L.B. conceived and designed the research project. R.J. performed all analyses and prepared the figures. M.L.B. supervised the research. R.J. and M.L.B. wrote the manuscript. Both authors reviewed the manuscript.

## Ethics Declarations

The authors declare no competing interests.

