## Supplementary material for "Meta-analysis reveals transcription factors and DNA binding domain variants associated with congenital heart defect and orofacial cleft": Suppl Figs + Suppl Tables 1, 2, 3 and 8

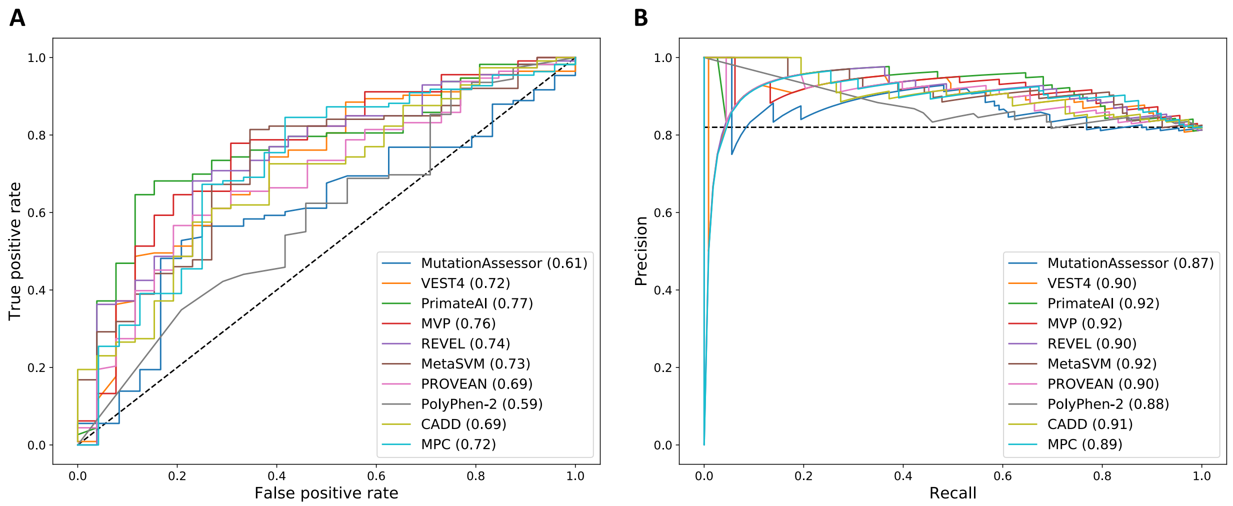


**Supplementary Figure 1. Performance metrics of missense variant prediction methods.** (**A**) Receiver operating characteristic curve for identifying CHD proband *de novo* variants. The values in parentheses are the area under the curve for each method. (**B**) Precision recall curve for identifying CHD proband *de novo* variants. The values in parentheses are the area under the curve for each method.

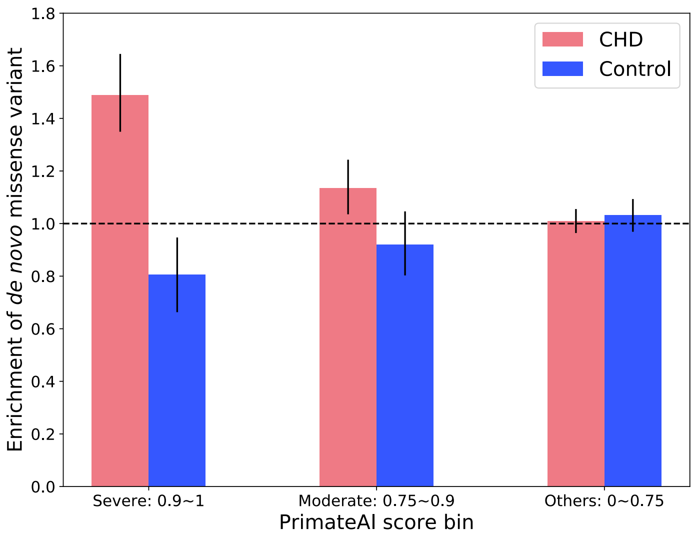


**Supplementary Figure 2. Enrichment of *de novo* missense variants for PrimateAI score ranges.** The enrichment is derived with respect to the expected number of *de novo* missense variants in cohorts of the same size as CHD trios (n = 3,835) and control trios (n = 2,179). The error bars are 95 % bootstrap confidence intervals.

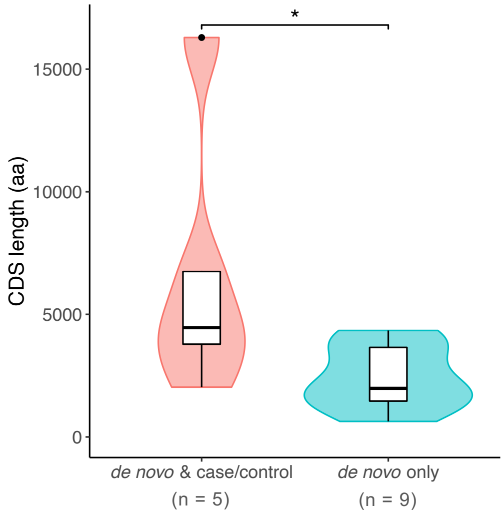


**Supplementary Figure 3. Coding sequence length of significant OC genes by discovery model.** Distribution of coding sequence length for the significant genes unique to the "*de novo* & case/control" model and "*de novo* only" model. The number of genes is labeled below each category. CDS, coding sequence; aa, amino acid. *: $p < 0.05$, one-sided Wilcoxon rank-sum test.

**Supplementary Table 1. Number of trios for CHD, OC, and unaffected children used for *de novo* variants.**

PCGC: Pediatric Cardiac Genomics Consortium, PHDN: Pediatric Heart Network, DDD: Deciphering Developmental Disorders

| Publication / dbGaP ID | Number of trios | Comments |
|---|---|---|
| **Congenital heart defect** | | |
| Jin *et al*. (1) | 2,645 | PCGC + PHN |
| Sifrim *et al*. (2) | 1,039 | DDD & others (discarded 326 also in Jin *et al*. (1)) |
| Richter *et al*. (3) | 151 | Kids First (discarded 612 also in Jin *et al*. (1)) |
| **Total** | **3,835** | |
| **Orofacial cleft** | | |
| Bishop *et al*. (4) | 757 | 374 European, 267 Colombian, and 116 Taiwanese |
| phs002595 | 351 | Family trios from the Philippines |
| phs001997 | 133 | African Craniofacial Anomalies Network (AfriCRAN) |
| Wilson *et al*. (5) | 603 | Deciphering Developmental Disorders |
| **Total** | **1,844** | |
| **Unaffected children** | | |
| Satterstrom *et al*.(6) | 2,165 | Unaffected sibling from families in an autism cohort |
| **Total** | **2,165** | |

**Supplementary Table 2. List of 225 Human CHD genes.**

The list was accessed at the Seidman Lab website (7) on 11/29/2023.

| | | | | | | | | |
|---|---|---|---|---|---|---|---|---|
| ABCC9 | BBS8 | COL3A1 | EOGT | GPC6 | MAP2K2 | OFD1 | RPS17 | SOX2 |
| ABCD3 | BBS9 | COL5A1 | EP300 | HAND1 | MED12 | PEX1 | RPS19 | SOX9 |
| ACTB | BCOR | COL5A2 | ESCO2 | HCCS | MED13L | PEX13 | RPS24 | STAMBP |
| ACVR2B | BRAF1 | COX7B | EVC | HOXA1 | MEGF8 | PHGDH | RPS26 | STRA6 |
| ADAMTS10 | CACNA1C | CREBBP | EVC2 | HRAS | MGP | PITX2 | RPS7 | TAB2 |
| ADNP | CBL | CRELD1 | FBN1 | IFT122 | MID1 | PKD1 | RSPH4A | TBX1 |
| ANKRD11 | CCDC103 | DDX11 | FBN2 | IFT140 | MKS1 | PKD2 | RSPH9 | TBX20 |
| ARHGAP31 | CCDC114 | DGCR2 | FGF8 | IFT80 | MYH6 | PLOD1 | SALL1 | TBX3 |
| ARID1A | CCDC151 | DHCR7 | FGFR1 | INVS | NEK1 | PQBP1 | SEMA3E | TBX5 |
| ARID1B | CCDC39 | DLL4 | FIG4 | IRX5 | NF1 | PTEN | SETBP1 | TCOF1 |
| ARL13B | CCDC40 | DNAAF1 | FKTN | JAG1 | NFATC1 | PTPN11 | SF3B4 | TFAP2B |
| ARMC4 | CD96 | DNAAF2 | FLNA | JBTS17 | NIPBL | RAB23 | SH3PXD2B | TGFBR1 |
| ASXL1 | CDK13 | DNAAF3 | FLT1 | KANSL1 | NKX2-5 | RAD21 | SHH | TGFBR2 |
| ATIC | CDKN1C | DNAH11 | FLT4 | KAT6A | NKX2-6 | RAF1 | SHOC2 | TLL1 |
| B3GALT6 | CEP290 | DNAH5 | FOXC1 | KAT6B | NME8 | RAI1 | SKI | TSC1 |
| BBS1 | CEP41 | DNAI1 | FOXC2 | KDR | NODAL | RBFOX2 | SMAD2 | TSC2 |
| BBS10 | CEP57 | DNAI2 | FOXF1 | KIF7 | NOTCH1 | RBM10 | SMAD3 | TTC21B |
| BBS11 | CFC1 | DNAL1 | FTO | KMT2A | NOTCH2 | RBM8A | SMAD4 | TWIST1 |
| BBS12 | CHD4 | DOCK6 | GATA4 | KMT2D | NPHP3 | RIT1 | SMAD6 | UBR1 |
| BBS2 | CHD7 | DYNC2H1 | GATA5 | KRAS | NPHP4 | ROR2 | SMARCA4 | WDR19 |
| BBS3 | CHST3 | DYXC1 | GATA6 | LBR | NPHP9 | RPGRIP1L | SMARCB1 | WDR35 |
| BBS4 | CITED2 | ECE1 | GDF1 | LEFTY2 | NR2F2 | RPL11 | SMARCE1 | WDR60 |
| BBS5 | COL1A1 | EFTUD2 | GJA1 | LRP2 | NRAS | RPL35A | SMC3 | ZEB2 |
| BBS6 | COL1A2 | EHMT1 | GLI3 | LTBP4 | NSD1 | RPL5 | SMS | ZFPM2 |
| BBS7 | COL2A1 | ELN | GPC3 | MAP2K1 | NSDHL | RPS10 | SOS1 | ZIC3 |

**Supplementary Table 3. List of 148 Human OC genes.**

The list was accessed at the National Health Service Genomic Medicine Service gene panels (8) for Clefting version 4.0 (labeled as Green for the highest level of confidence).

| | | | | | | | |
|---|---|---|---|---|---|---|---|
| *ACTB* | *COL11A2* | *EIF2S3* | *HDAC8* | *MBTPS2* | *PLCB4* | *SKI* | *TGFBR1* |
| *ACTG1* | *COL2A1* | *EIF4A3* | *HYAL2* | *MED12* | *POLR1B* | *SLC26A2* | *TGFBR2* |
| *AMER1* | *COL9A1* | *EOGT* | *HYLS1* | *MED25* | *POLR1C* | *SMAD3* | *TMCO1* |
| *ANKRD11* | *COLEC10* | *EPG5* | *ICK* | *MEIS2* | *POLR1D* | *SMAD4* | *TP63* |
| *ARHGAP29* | *COLEC11* | *ESCO2* | *IFT140* | *MID1* | *PORCN* | *SMC1A* | *TRAPPC9* |
| *ARHGAP31* | *CTCF* | *EYA1* | *IFT172* | *MKS1* | *PTCH1* | *SMC3* | *TRIM37* |
| *ASXL1* | *CTNND1* | *FAM20C* | *IFT80* | *MSX1* | *RBM10* | *SMS* | *TUBB* |
| *B3GLCT* | *DHCR7* | *FGD1* | *IMPAD1* | *MYMK* | *ROR2* | *SNRPB* | *TXNL4A* |
| *BCOR* | *DHODH* | *FGFR1* | *IRF6* | *NECTIN1* | *RPL5* | *SON* | *USP9X* |
| *BMP2* | *DLL4* | *FGFR2* | *KAT6A* | *NEDD4L* | *RPS26* | *SOX9* | *WNT5A* |
| *C2CD3* | *DOCK6* | *FLNA* | *KCNJ2* | *NEK1* | *SALL4* | *SPECC1L* | *XYLT1* |
| *C5orf42* | *DVL1* | *FLNB* | *KDM6A* | *NIPBL* | *SATB2* | *STAMBP* | *ZEB2* |
| *CC2D2A* | *DVL3* | *FOXC2* | *KIAA0586* | *NOTCH1* | *SCARF2* | *TBX22* | *ZIC2* |
| *CDH1* | *DYNC2H1* | *FRAS1* | *KIF1BP* | *OFD1* | *SF3B2* | *TCOF1* | *ZIC3* |
| *CDKN1C* | *DYNC2LI1* | *GDF11* | *KIF7* | *PAX3* | *SF3B4* | *TCTN3* | *ZSWIM6* |
| *CHD7* | *EBP* | *GJA1* | *KMT2D* | *PHF8* | *SHH* | *TELO2* | |
| *CHRNG* | *EDNRA* | *GLI3* | *MAP3K7* | *PIEZO2* | *SIX1* | *TFAP2A* | |
| *CHST14* | *EFNB1* | *GPC3* | *MAPRE2* | *PIGN* | *SIX3* | *TGDS* | |
| *COL11A1* | *EFTUD2* | *GRHL3* | *MASP1* | *PIGV* | *SIX5* | *TGFB3* | |

**Supplementary Table 4 (See Excel file) TADA results for CHD**

**Supplementary Table 5 (See Excel file) TADA results for OC**

**Supplementary Table 6 (See Excel file) *De novo* missense variants in CHD genes in CHD cohorts**

**Supplementary Table 7 (See Excel file) *De novo* missense variants in OC genes in OC cohorts**

**Supplementary Table 8 TF DBD Pfam domains**

| Domain name | Pfam ID |
|---|---|
| TF_AP-2 | PF03299 |
| ARID | PF01388 |
| AT_hook | PF02178 |
| zf-BED | PF02892 |
| Basic | PF01586 |
| HLH | PF00010 |
| BrkDBD | PF09607 |
| bZIP_1 | PF00170 |
| bZIP_2 | PF07716 |
| bZIP_Maf | PF03131 |
| zf-C2H2 | PF00096 |
| CBFB_NFYA | PF02045 |
| zf-CCCH | PF00642 |
| CBFD_NFYB_HMF | PF00808 |
| CENP-B_N | PF04218 |
| CG-1 | PF03859 |
| CSD | PF00313 |
| CUT | PF02376 |
| zf-C2HC | PF01530 |
| zf-CXXC | PF02008 |
| DM | PF00751 |
| E2F_TDP | PF02319 |
| COE1_DBD | PF16422 |
| COE1_HLH | PF16423 |
| Ets | PF00178 |
| FLYWCH | PF04500 |
| Forkhead | PF00250 |
| GATA | PF00320 |
| GCM | PF03615 |
| CP2 | PF04516 |
| GTF2I | PF02946 |
| HMG_box | PF00505 |
| Homeodomain | PF00046 |

(Continued)

**Supplementary Table 8** (Continued)

| Domain name | Pfam ID |
|---|---|
| HSF_DNA-bind | PF00447 |
| IRF | PF00605 |
| IRF-3 | PF10401 |
| SRF-TF | PF00319 |
| MBD | PF01429 |
| mTERF | PF02536 |
| Myb_DNA-binding | PF00249 |
| Myb_DNA-bind_4 | PF13837 |
| Myb_DNA-bind_5 | PF13873 |
| NDT80_PhoG | PF05224 |
| zf-NF-X1 | PF01422 |
| zf-C4 | PF00105 |
| zf-CCHC | PF00098 |
| P53 | PF00870 |
| PAX | PF00292 |
| Pou | PF00157 |
| Prox1 | PF05044 |
| HTH_psq | PF05225 |
| RHD_DNA_bind | PF00554 |
| RFX_DNA_binding | PF02257 |
| Runt | PF00853 |
| SAND | PF01342 |
| MH1 | PF03165 |
| STAT_bind | PF02864 |
| TBX | PF12598 |
| T-box | PF00907 |
| TBP | PF00352 |
| TCF | PF03638 |
| TEA | PF01285 |
| THAP | PF05485 |

**Supplementary Table 9 (See Excel file) *De novo* TF DBD missense variants in the CHD and OC cohorts**
